# Quantitative CTA-Based Carotid Plaque Composition Across Stroke Etiologies and Residual Vascular Risk

**DOI:** 10.64898/2026.10.28.26366685

**Authors:** Yingle Li, Luka Živković, Roya Batool, Abdalla Marei, Paul Reidler, Anna Kopczak, Luca Saba, Nikolaos Tsilimparis, Amir Ahmadi, Robert A. Pelberg, DEMDAS Study Group, Marios K. Georgakis

**Author notes:** **Corresponding author information:** Marios K. Georgakis, MD, PhD, Institute for Stroke and Dementia Research (ISD), LMU University Hospital, Feodor-Lynen-Straße 17, 81377 Munich, Germany.

## Abstract

**Background:** Carotid plaque vulnerability features may provide prognostic information beyond luminal stenosis, but the value of quantitative plaque composition on CTA remains unclear.

**Objectives:** We investigated whether plaque composition analysis in routine CTA in patients with ischemic stroke or TIA can identify high-risk plaques across etiologies, particularly among strokes of undetermined etiology and predict residual vascular risk.

**Methods:** We analyzed carotid CTAs from 314 patients with acute ischemic stroke or TIA. CTA-derived plaque composition was quantified using commercially available software and compared between ipsilateral and contralateral plaques across etiology groups. Associations of plaque characteristics with subsequent vascular events were assessed using Cox regression models.

**Results:** Among patients with ischemic events of undetermined etiology, ipsilateral plaques showed significant enrichment for greater LRNC volume (ratio, 1.97; 95% CI, 1.22–3.18), an association that persisted after adjustment for wall volume (ratio, 1.51; 95% CI, 1.15–1.99). During a median 3-year follow-up, larger LRNC volume was associated with subsequent vascular events in patients with ischemic stroke of undetermined etiology (HR, 1.96; 95% CI, 1.12–3.43), as were calcification, noncalcified matrix, and total wall volumes. Associations for individual plaque components attenuated after adjustment for wall volume.

**Conclusions:** CTA-derived LRNC is enriched in carotid plaques ipsilateral to ischemic events of undetermined etiology and is associated with future vascular risk. Quantitative CTA-based plaque assessment may provide complementary information for etiologic evaluation and vascular risk stratification.

## Introduction

Carotid atherosclerosis is a major cause of ischemic stroke and transient ischemic attack (TIA), yet clinical assessment of carotid plaques continues to rely predominantly on the degree of luminal stenosis.^1,2^ Specifically, only plaques leading to >50% stenosis of the internal carotid artery are considered to be the underlying cause of an ipsilateral stroke or TIA.^3^ However, luminal stenosis provides only a limited representation of plaque biology and vulnerability.^4^ Plaques producing a similar degree of narrowing may differ substantially in total wall burden, lipid-rich necrotic core (LRNC), calcification, noncalcified matrix, remodeling, and other features related to plaque vulnerability. These features have been most extensively characterized *in vivo* using high-resolution multi-contrast carotid MRI,^5,6^ which offers sufficient resolution for assessing plaque composition.

However, dedicated carotid plaque MRI requires additional acquisition time, specialized imaging protocols, and expert post-processing, and is therefore not routinely performed during the acute evaluation of stroke patients. In contrast, computed tomography angiography (CTA) is routinely acquired as part of the acute stroke work-up,^7^ making CTA-based quantitative assessment of the vessel wall and plaque composition an attractive approach to provide clinically relevant information beyond conventional lumen-based evaluation without requiring an additional examination.^8,9^ Despite the traditional view that CTA lacks sufficient spatial and contrast resolution to characterize carotid plaque composition, recent studies with histopathological validation suggest that several plaque components can be reliably quantified using carotid CTA.^10^ Particularly, CTA appears capable of identifying and quantifying calcified and non-calcified plaque components, as well as more specific high-risk features such as LRNC.^10^

This issue is particularly relevant in patients with ischemic events of undetermined etiology, in whom the underlying mechanism remains uncertain. Previous MRI studies have reported an excess of high-risk features in ipsilateral non-stenotic carotid plaques in patients with undetermined stroke.^11^ However, it remains unclear whether quantitative plaque composition extracted from routinely acquired acute-stroke CTA shows distinct side-specific patterns across stroke etiologies, and provides prognostic information on subsequent vascular risk, particularly in patients with stroke of undetermined etiology.

To address these gaps, we applied a commercially available quantitative plaque-analysis approach to routinely acquired acute-stroke CTA across four cohorts. We first investigated side-specific differences in carotid plaque composition across stroke etiologies, with particular focus on patients with stroke of undetermined etiology. We then evaluated whether patient-level carotid plaque burden and composition were associated with subsequent vascular events.

## Methods

### Study cohorts and inclusion criteria

For the purposes of this study, we leveraged data from four studies of stroke or TIA patients at the LMU University Hospital in Munich, Germany. The DEDEMAS (recruitment 2011–2013, single-center)^12,13^ and DEMDAS (recruitment 2013–2018, multicenter across six tertiary stroke centers in Germany)^14,15^ studies are observational cohorts designed to investigate predictors of cognitive decline after acute stroke; in the current analysis, only patients recruited at the LMU University Hospital were included. PROSCIS-M (recruitment 2011–2023)^16^ is the Munich arm of a prospective cohort designed to identify predictors of long-term vascular outcomes after first-ever ischemic stroke. AtherOMICS (recruitment 2022–2026)^17^ is a prospective study and a biobank integrating vascular imaging, blood, plaque tissue, and multi-omic profiling in patients undergoing carotid or femoral endarterectomy. Details regarding the study design of each source study have been previously published^12–17^ and are also summarized in the Supplementary Methods (Section S1.1).

For the present analysis, eligible participants were aged ≥18 years, had been recruited due to an ischemic stroke or TIA in one of the abovementioned studies at the LMU University Hospital site, had cervical CTA demonstrating the presence of carotid plaque suitable for quantitative analysis, and had sufficient clinical information to confirm the ischemic event and classify its etiology and vascular territory. Participants were excluded if they had a non-ischemic event, a hemorrhagic stroke, lacked CTA, or had CTA that did not fulfill the criteria required for reliable quantitative plaque analysis (section thickness >1.5 mm or insufficient image quality, e.g., due to motion artifacts or artifacts from dental prostheses). Additional exclusions included technical processing failures and anatomical or acquisition-related limitations that precluded reliable plaque segmentation. AtherOMICS participants were excluded from the longitudinal analysis because follow-up data were not available for this cohort.

All participants provided written informed consent for participation in the studies. All studies received ethical approval from the Ethics Committee of the Medical Faculty of LMU Munich: DEDEMAS/DEMDAS (approval numbers 035–11 and 201–13), PROSCIS-M (approval number 366–10), and AtherOMICS (approval numbers 121–09 and 22–0135). The retrospective analysis of the CTA dataset was also approved by the Ethics Committee of the Medical Faculty of LMU Munich (23–0772).

### CTA acquisition and plaque feature quantification

Cervical CTAs were acquired during routine acute stroke imaging using multidetector CT scanners. Only examinations with reconstructed section thickness ≤1.5 mm and sufficient image quality were included. Detailed acquisition parameters are provided in the Supplementary Methods (Section S1.2). CTA datasets were analyzed using the PlaqueIQ™ software (Elucid Inc., Boston, MA, USA),^10^ which segmented the luminal and outer vessel wall boundaries and quantified lumen volume, wall volume, and plaque-component volumes, including LRNC, calcification, and noncalcified matrix. Wall volume was defined as the three-dimensional volume of tissue between the luminal and outer vessel wall boundaries within the analyzed carotid region, encompassing plaque tissue as well as any non-plaque wall tissue present within these boundaries. Plaque analysis covered the distal common carotid artery to the proximal internal carotid artery.

### Ischemic event etiology and vascular territory

All patients underwent routine diagnostic evaluation for acute ischemic stroke, including clinical assessment, laboratory testing, electrocardiography, cardiac monitoring, vascular imaging, brain imaging, and echocardiography when clinically indicated. Stroke etiology was classified according to the Trial of ORG 10172 in Acute Stroke Treatment (TOAST) criteria using all available clinical and diagnostic information, independently of the quantitative carotid plaque measurements.^3^ For the purposes of the current study, index events were categorized as carotid atherosclerosis-related LAA etiology, other determined etiology, or undetermined etiology, the latter including incomplete diagnostic workup, undetermined etiology despite complete workup, and competing etiologies. Events of other determined etiologies comprised events related to intracranial large-artery atherosclerosis, large artery atherosclerotic events in the posterior circulation, cardioembolism, small-vessel occlusion, and stroke of other determined etiology in the TOAST 4 category. For simplicity, the three categories are described as carotid LAA, other determined etiology, and undetermined etiology throughout. The side of the index event was assigned according to the vascular territory of the acute infarct on brain MRI, supported by clinical presentation and vascular imaging. Carotid plaques were classified as ipsilateral, if ipsilateral to the ischemic event, or contralateral if not. For the purposes of the primary laterality analysis, both carotid plaques were coded as contralateral in patients with 死哈 circulation events and as ipsilateral in those with bilateral index events; both groups were excluded in sensitivity analyses.

### Assessment of future vascular events

In DEDEMAS^12,13^/DEMDAS^14,15^, comprehensive in-person assessments were scheduled at 6, 12, 36, and 60 months, with telephone assessments at 3, 24, and 48 months. In PROSCIS-M^16^, follow-up assessments were conducted by telephone at 3, 12, 24, and 36 months. Reported outcome events were verified using medical records, imaging, and documentation from treating physicians or hospitals. The composite endpoint comprised recurrent stroke, transient ischemic attack, myocardial infarction, angina pectoris, coronary artery disease, or peripheral artery disease during follow-up. Follow-up extended from CTA to the first endpoint or last available assessment. Cohort-specific follow-up schedules are provided in the Supplementary Methods (Section S1.3).

### Statistical analysis

Analyses addressed two primary questions: whether CTA-derived carotid plaque characteristics differed between plaques ipsilateral to the ischemic event vs. other plaques, and whether baseline plaque characteristics were associated with subsequent vascular events. All analyses were performed separately within the carotid LAA, other determined etiology, and undetermined etiology groups. Because plaque-volume measures were right-skewed, they were transformed using log10(value + 1) before modeling.

For cross-sectional analyses, the carotid plaque was the unit of analysis. Each plaque characteristic was modeled using age- and sex-adjusted linear mixed-effects models with a participant-specific random intercept. Estimates were back-transformed and reported as ipsilateral-to-contralateral geometric mean ratios with 95% confidence intervals. Additional adjusted and sensitivity analyses are described in the Supplementary Methods (Section S2.1).

For the longitudinal analysis, the participant was the unit of analysis. Bilateral total plaque measures were calculated across evaluable carotid plaques, transformed, standardized within each etiology group, and entered separately into age- and sex-adjusted Cox proportional hazards models. Hazard ratios with 95% confidence intervals were reported per 1-standard-deviation increase. Additional adjusted and sensitivity analyses, including alternative wall-volume summaries, are described in the Supplementary Methods (Section S2.2).

False discovery rate (FDR) was controlled using the Benjamini–Hochberg procedure, with correction performed separately within each prespecified stroke-etiology group.^18^ Details of the correction families are provided in the Supplementary Methods (Section S2.3).

## Results

### Study workflow and baseline characteristics of study population

Carotid plaque composition could be assessed in CTAs of a total of 372 patients. After excluding asymptomatic patients without an acute ischemic event (n = 51 from the AtherOMICS biobank), and those with an intracerebral hemorrhage (n = 7), 314 patients could be included in the final analysis (Figure 1). Among them, 127 patients had an ischemic event classified as of carotid LAA origin, 87 as of another determined etiology, and 100 as of an undetermined etiology. Within the undetermined etiology group, 16 patients (16.0%) had an incomplete diagnostic workup, 67 (67.0%) had no identifiable etiology despite a complete workup, and 17 (17.0%) had competing potential etiologies. Overall, 625 carotid plaques were analyzed including 253 plaques in the carotid LAA group, 173 in the other determined etiology group, and 199 in the undetermined etiology.

**Figure 1.**
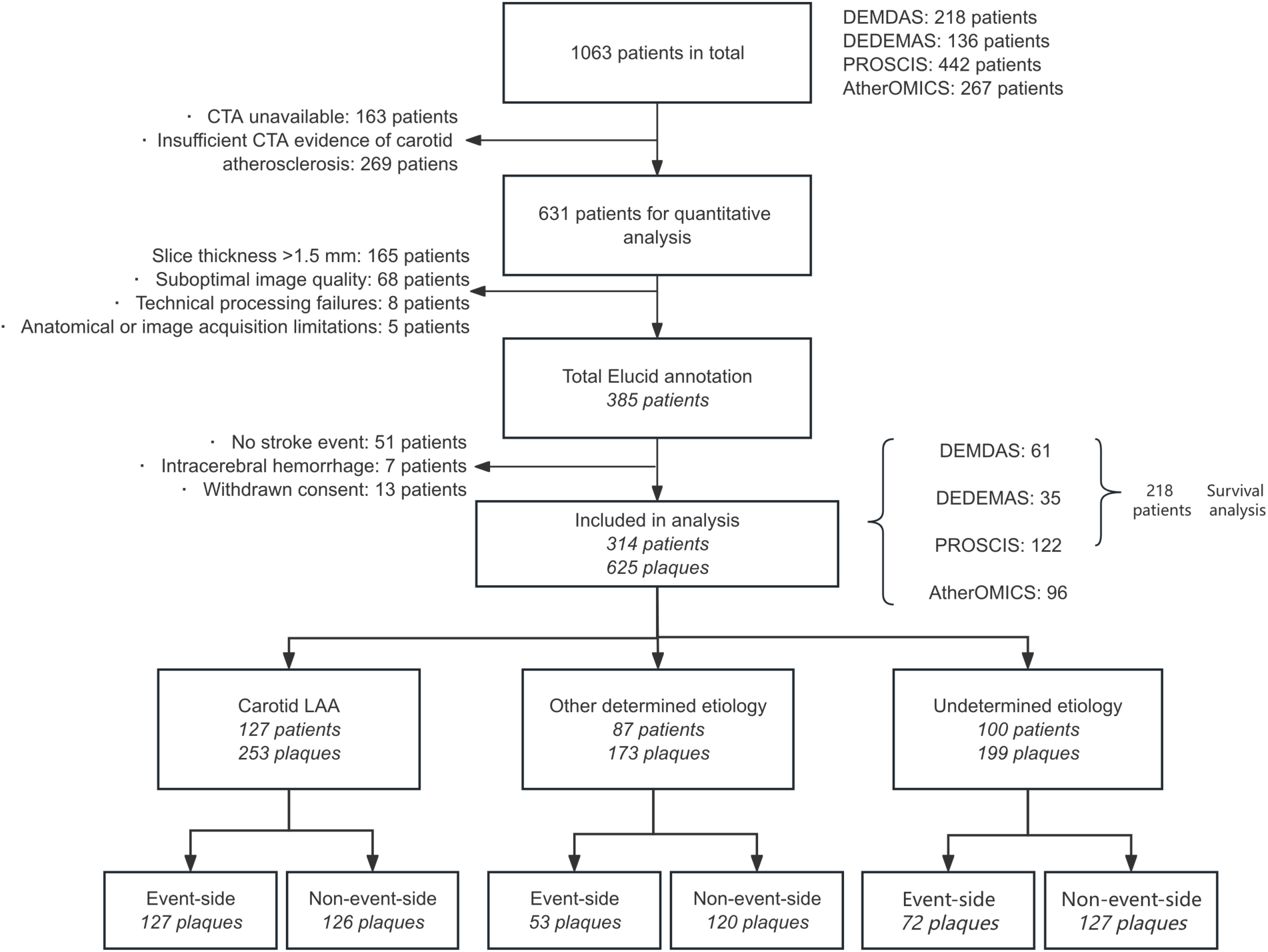
Study population and analytical workflow. Flow diagram illustrating participant selection, stroke etiology classification, and allocation to cross-sectional and longitudinal analyses. The cross-sectional analysis included 314 patients and 625 carotid plaques, with plaques classified as ipsilateral or contralateral to the qualifying ischemic event. The longitudinal analysis included 218 patients with available follow-up data from DEDEMAS, DEMDAS, and PROSCIS-M. CTA = computed tomography angiography; LAA = large-artery atherosclerosis.

Baseline characteristics according to ischemic event etiology are summarized in Table 1, with cohort-specific characteristics presented in Supplementary Table 1. Patients with carotid LAA were older, more often male, and had a greater vascular risk burden than those with other determined or undetermined etiologies. Overall index-event severity was mild (median NIHSS score, 2). Included participants were older and more frequently had previous stroke and diabetes than excluded participants (Supplementary Table 2).

**Table 1.** Baseline Characteristics of Included Patients According to the Etiology of Index Ischemic Event.

| Variables | Overall<br>n = 314 | Carotid<br>LAA<br>n = 127 | Other<br>determined<br>n = 87 | Undetermined<br>n = 100 | P-value |
| --- | --- | --- | --- | --- | --- |
| Age, years, mean ± SD | 73.0 ± 9.7 | 74.1 ± 9.0 | 73.8 ± 10.2 | 71.0 ± 9.8 | 0.038 |
| Female sex, n (%) | 165 (52.5) | 45 (35.4) | 53 (60.9) | 67 (67.0) | <0.001 |
| BMI, kg/m<sup>2</sup>, mean ± SD | 26.5 ± 4.1 | 26.6 ± 4.1 | 26.4 ± 4.1 | 26.4 ± 4.0 | 0.877 |
| Hypertension, n (%) | 231 (73.8) | 89 (70.1) | 65 (75.6) | 77 (77.0) | 0.459 |
| Diabetes mellitus, n (%) | 72 (22.9) | 33 (26.0) | 18 (20.7) | 21 (21.0) | 0.564 |
| Dyslipidemia, n (%) | 135 (44.4) | 69 (59.0) | 28 (32.2) | 38 (38.0) | <0.001 |
| Ever smoking, n (%) | 152 (48.4) | 76 (59.8) | 32 (36.8) | 44 (44.0) | 0.003 |
| Coronary artery disease, n (%) | 64 (20.6) | 33 (26.8) | 14 (16.1) | 17 (17.0) | 0.101 |
| Previous stroke, n (%) | 45 (14.8) | 33 (28.2) | 3 (3.4) | 9 (9.0) | <0.001 |
| Peripheral artery disease, n (%) | 26 (8.5) | 18 (14.9) | 3 (3.6) | 5 (5.0) | 0.005 |
| Family history of CVD, n (%) | 163 (53.8) | 60 (51.3) | 48 (55.8) | 55 (55.0) | 0.801 |
| Baseline NIHSS, median (IQR) | 2.0 (1.0-4.0) | 1.0 (0.2-3.8) | 2.0 (1.0-4.8) | 3.0 (1.0-5.5) | <0.001 |

Continuous variables are shown as mean ± SD when approximately following a normal distribution and as median (IQR) when distributions were skewed; categorical variables are shown as n (%). P-values represent permutation-based tests of overall differences across the three groups. Summary statistics were calculated using available data; denominators may vary because of missing values in some of the presented variables. BMI, body mass index; IQR, interquartile range; LAA, large-artery atherosclerosis; CVD, cardiovascular disease; NIHSS, National Institutes of Health Stroke Scale; SD, standard deviation.

### Ipsilateral enrichment of carotid plaque composition differs across stroke etiologies

CTA-derived plaque volumes stratified by plaque side and stroke etiology are presented in Figure 2. We next compared ipsilateral and contralateral carotid plaque composition across ischemic event etiologies (Figure 3A). Ipsilateral plaque enrichment differed markedly across etiologies. Among patients with carotid LAA-related ischemic events, ipsilateral plaques demonstrated significantly greater overall wall volume and smaller lumen volume than contralateral plaques. They also contained larger volumes of all quantified plaque components, including calcification, lipid-rich necrotic core, and non-calcified matrix (all FDR < 0.001). The strongest association was observed for LRNC volume (Ratio, 3.68, 95% CI 2.62–5.16; FDR<0.001), which remained significant across additional adjusted models (Supplementary Table 3). In contrast, patients with ischemic events attributed to other determined etiologies showed no evidence of ipsilateral enrichment for any plaque characteristic, supporting the specificity of these findings to carotid atherosclerosis-related events.

**Figure 2.**
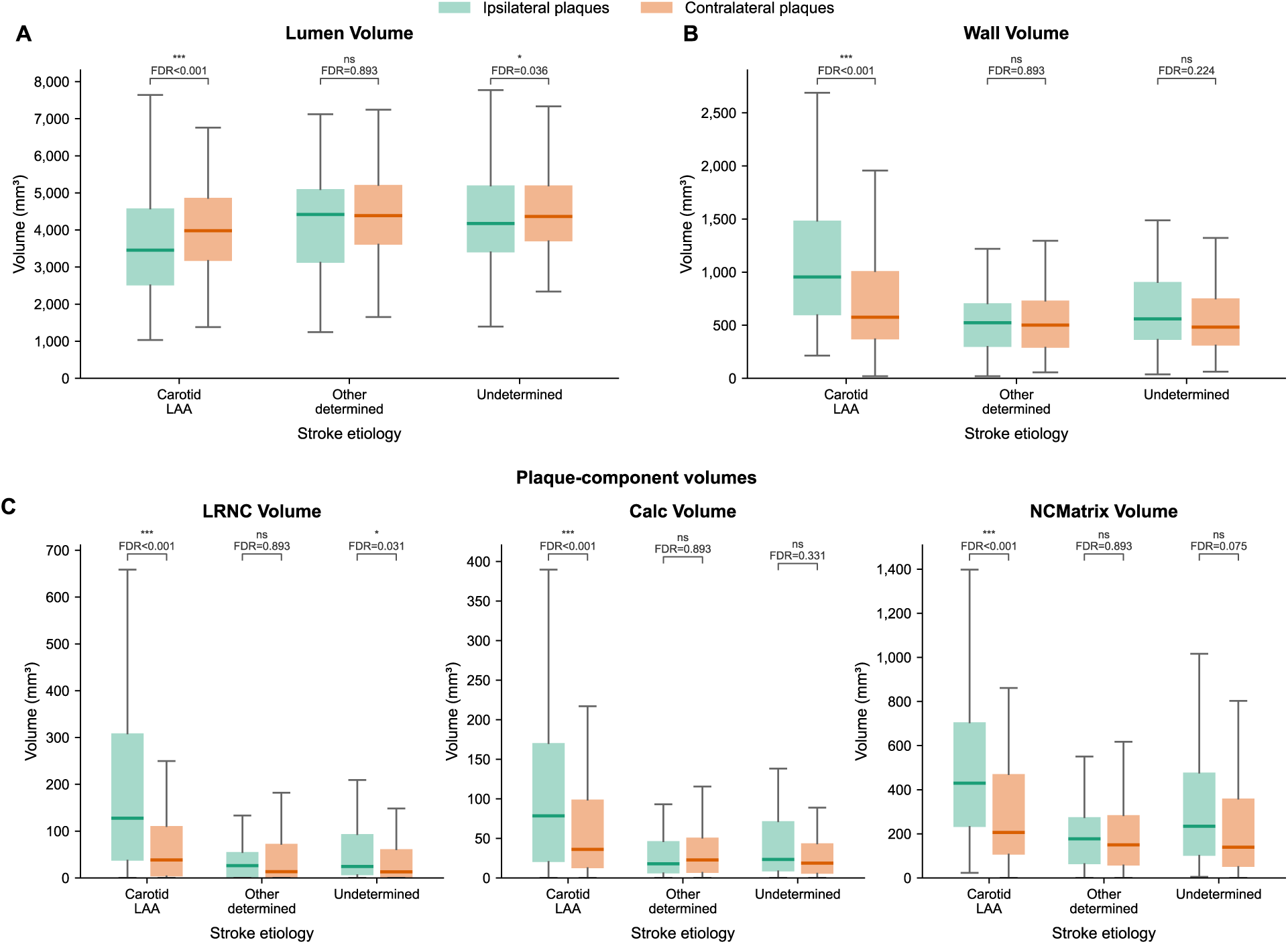
Plaque-component Volumes According to Plaque Side and Stroke Etiology. (A) Lumen volume and (B) wall volume in ipsilateral and contralateral carotid plaques, shown separately for the Carotid LAA, other determined, and undetermined etiology groups. (C) LRNC, CALC, and NCMatrix volumes across stroke etiologies, shown separately for each plaque component. Boxes represent the interquartile range, and horizontal lines indicate medians. All volumes are expressed in mm³. LRNC indicates lipid-rich necrotic core; CALC, calcification; and NCMatrix, noncalcified matrix.

**Figure 3.**
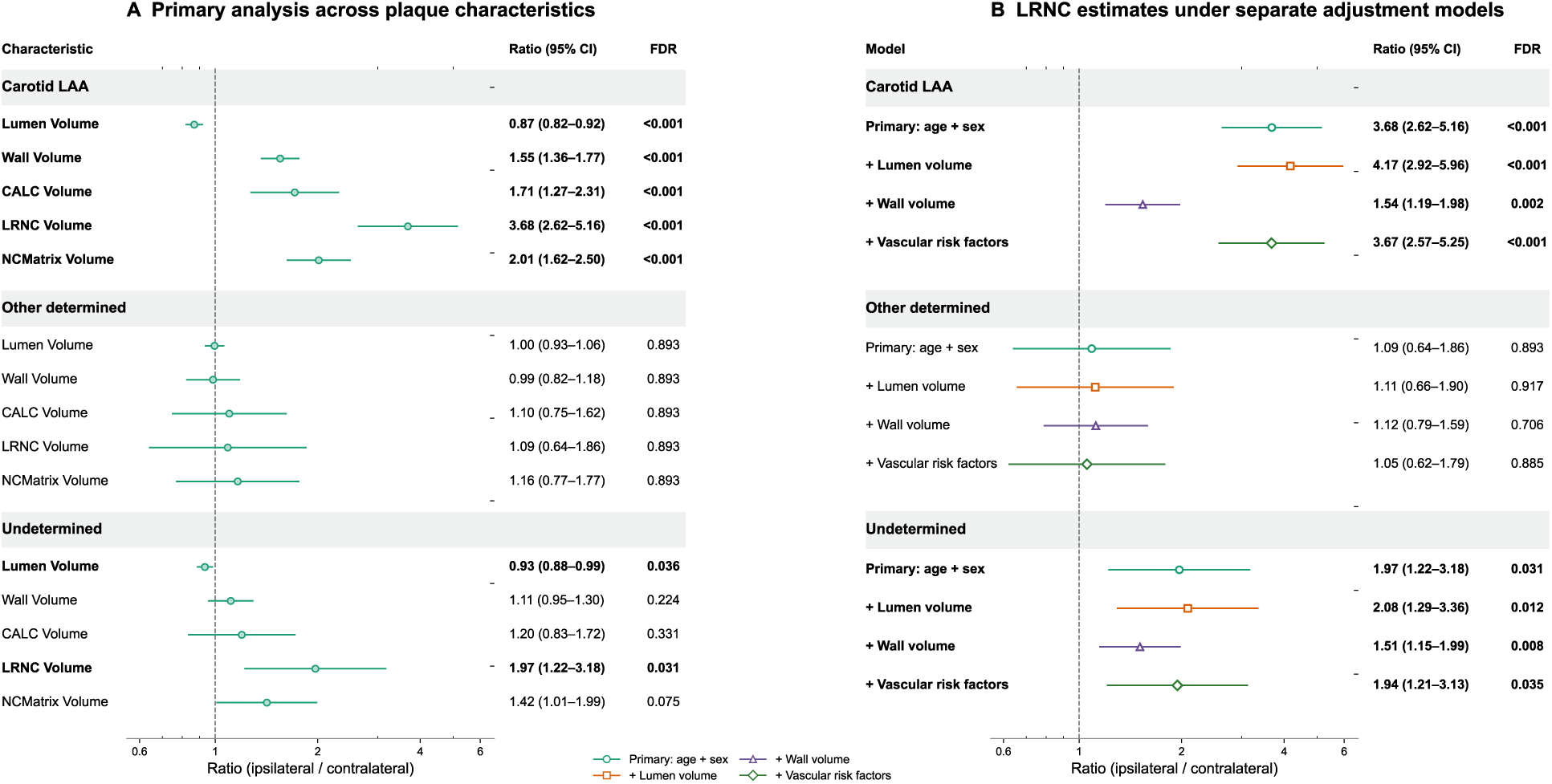
Ipsilateral Plaque Enrichment Across Stroke Etiologies. (A) Ipsilateral-to-contralateral ratios for five plaque characteristics from age- and sex-adjusted linear mixed-effects models. (B) LRNC ratios from the primary model and three separately adjusted models additionally accounting for lumen volume, wall volume, or vascular risk factors (hypertension, diabetes, dyslipidemia, smoking, and BMI). Points and horizontal lines represent back-transformed ratios and 95% confidence intervals. Ratios greater than 1 indicate higher values in ipsilateral plaques. The dashed vertical line at 1 indicates no side-specific difference. Bold values indicate FDR-adjusted P < 0.05. BMI, body mass index; CALC, calcification; CI, confidence interval; FDR, false discovery rate; LAA, large-artery atherosclerosis; LRNC, lipid-rich necrotic core; NCMatrix, noncalcified matrix.

Among patients with stroke of undetermined etiology, ipsilateral plaques exhibited a distinct pattern. While overall wall volume and calcification volume did not differ significantly between sides, ipsilateral plaques were enriched for LRNC (Ratio 1.97, 95% CI 1.22–3.18; FDR =0.031) and had smaller lumen volume (Ratio 0.93, 95% CI 0.88–0.99; FDR =0.036). Importantly, the ipsilateral enrichment of LRNC persisted after additional adjustment for wall volume (ratio, 1.51; 95% CI, 1.15–1.99; FDR = 0.008), suggesting a difference in plaque composition beyond overall wall burden. This association also remained significant in separate models additionally adjusted for lumen volume (ratio, 2.08; 95% CI, 1.29–3.36; FDR = 0.012) or conventional vascular risk factors (ratio, 1.94; 95% CI, 1.21–3.13; FDR = 0.035) (Figure 3B; Supplementary Table 3). Sensitivity analyses yielded directionally consistent LRNC estimates in the paired analysis and after excluding patients with posterior-circulation or bilateral index-event patterns (Supplementary Table 4). Collectively, these findings suggest that CTA-derived LRNC identifies carotid plaques associated with the index ischemic events not only in carotid LAA-related events but also in a subset of patients with events of undetermined etiology.

### CTA-derived carotid plaque composition is associated with residual vascular risk in ischemic strokes of undetermined etiology

We next examined whether patient-level carotid plaque characteristics were associated with subsequent vascular events across stroke etiologies. The longitudinal analysis included 218 ischemic stroke patients, comprising 31 with carotid LAA-related strokes, 87 with stroke of other determined etiologies, and 100 with stroke of undetermined etiology. During a median follow-up of 35.6 months (IQR, 22.9–47.8), the composite vascular endpoint occurred in 45 patients (20.6%), including 10 of 31 patients with carotid LAA-related stroke (32.3%), 15 of 87 patients with ischemic strokes of other determined etiologies (17.2%), and 20 of 100 patients with stroke of undetermined etiology (20.0%). The distribution of first follow-up events by stroke etiology is presented in Supplementary Table 9.

In etiology-stratified Cox regression analyses, no plaque burden or plaque composition measure was significantly associated with the composite vascular endpoint among patients with carotid LAA-related strokes or ischemic strokes of other determined etiologies after FDR correction. In contrast, among patients with ischemic stroke of undetermined etiology, greater bilateral total wall volume, calcification volume, LRNC volume, and noncalcified matrix volume were associated with a higher risk of the composite vascular endpoint after FDR correction (Figure 4a; Supplementary Table 5). For total wall volume, the age-and sex-adjusted HR was 2.21 (95% CI, 1.35–3.61; P = 0.002; FDR = 0.008) (Figure 4a; Supplementary Table 5). When plaque-composition measures were evaluated, their associations were attenuated after additional adjustment for total wall volume. In separate models evaluating wall volume, the association remained robust after adjustment for total lumen volume and conventional vascular risk factors (Supplementary Table 6). In sensitivity analyses that either simultaneously included ipsilateral and contralateral plaque measures or excluded patients with posterior-circulation or bilateral index events, effect estimates in the undetermined etiology group were generally similar in direction and magnitude to those of the primary analyses, although confidence intervals were wider (Supplementary Table 7). In the alternative wall-volume analysis, all four wall-volume summaries were associated with the composite vascular endpoint after FDR correction: bilateral total, maximum-side, ipsilateral, and contralateral wall volume (Figure 4b; Supplementary Table 8).

**Figure 4.**
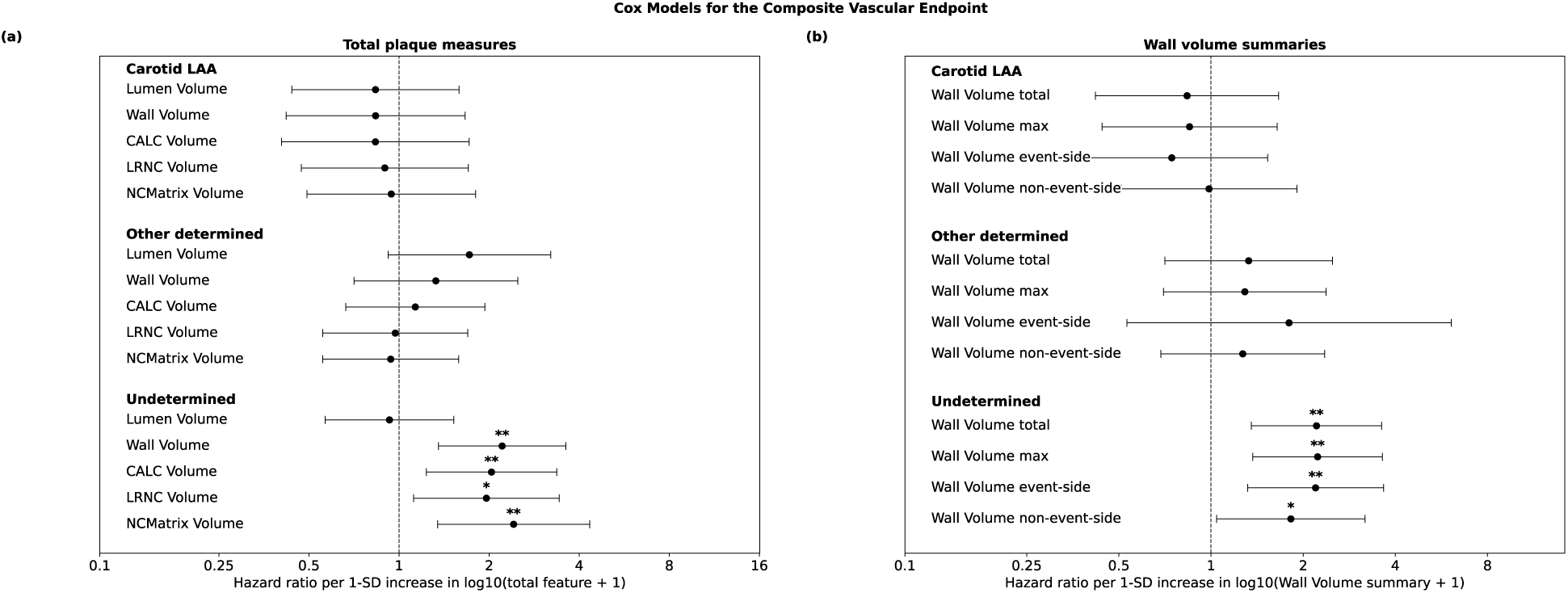
Carotid Plaque Features and Subsequent Vascular Events Across Stroke Etiologies. Age- and sex-adjusted HRs and 95% CIs per 1-SD increase are shown for (A) bilateral total plaque measures and (B) alternative wall-volume measures. Stars indicate FDR-adjusted significance.

## Discussion

The main findings are summarized in the Central Illustration. In this pooled analysis of patients with ischemic stroke or TIA, we found that quantitative assessment of carotid plaque composition on routine CTA can reliably identify high-risk features with potential implications for etiology assessment and recurrence risk stratification. First, ipsilateral plaques in patients with events of undetermined etiology showed significant enrichment for greater LRNC volume, consistent with the pattern observed in carotid atherosclerosis-related ischemic events. Second, in the same group, greater plaque burden and larger LRNC, calcification, and noncalcified matrix volumes were associated with future vascular events over a median follow-up of three years.

We found selective enrichment of LRNC in carotid plaques ipsilateral to ischemic events of undetermined etiology. Although the association was more pronounced in carotid LAA in the age- and sex-adjusted analyses, the effect estimates were numerically similar after additional adjustment for wall volume (ratio, 1.54; 95% CI, 1.19–1.98 in carotid LAA; ratio, 1.51; 95% CI, 1.15–1.99 in undetermined etiology). This finding is consistent with a shared pattern of event-side LRNC enrichment across the two etiologic groups, which was not fully explained by differences in wall volume. In the undetermined etiology group, the association also remained significant in separate models accounting for lumen volume or conventional vascular risk factors. These findings support the potential value of quantitative LRNC assessment for characterizing carotid plaques associated with the presenting ischemic event, beyond measures of overall plaque burden, including in patients not meeting conventional stenosis-based criteria for carotid LAA. Previous studies have shown that non-stenotic carotid plaques with greater plaque thickness, complicated morphology, intraplaque hemorrhage, or other high-risk features occur more frequently ipsilateral to the infarct in patients with cryptogenic stroke or embolic stroke of undetermined source.^11,19–24^ Most of these studies, however, were based on high-resolution carotid MRI,^11,21,22^ a modality that is difficult to acquire routinely in all patients with ischemic events of unknown etiology. Previous studies assessing plaque characteristics on CTA reported a higher frequency of greater plaque thickness ipsilateral to cryptogenic events.^19,25–27^ Our findings extend these observations by showing that differences in plaque composition can also be quantitatively captured on routine CTA. While these ipsilateral differences cannot establish causality, as other occult embolic sources may coexist in this heterogeneous population,^28,29^ our findings suggest that quantitative CTA can detect relevant carotid atherosclerosis beyond stenosis-based definitions. This may have implications for the etiologic classification and subsequently secondary prevention strategies in ischemic events of undetermined etiology, which still account for >30% of ischemic strokes in clinical practice.^8^

Beyond associations with the index event, we found that high-risk plaque features, including LRNC, and higher plaque burden were also associated with future vascular events in patients with ischemic events of undetermined etiology. Notably, the associations of bilateral, ipsilateral, and contralateral wall volume with subsequent vascular events suggest that carotid plaque burden may capture a generalized atherosclerotic phenotype rather than risk arising solely from the index carotid plaque. The attenuation of associations for individual plaque components after adjustment for wall volume further suggests that overall atherosclerotic burden may be particularly relevant to long-term vascular risk. These associations were statistically significant in the undetermined etiology group and were not statistically significant in patients with carotid LAA, potentially reflecting plaque removal through carotid endarterectomy, which is recommended for eligible patients with carotid stenosis meeting the TOAST criteria for LAA. However, the longitudinal LAA analysis included only 31 patients with 10 events. The low number of subsequent events precluded meaningful comparisons of event subtypes, although recurrent stroke or TIA accounted for nearly half of the composite events (48.9%). Previous longitudinal studies have similarly linked MRI-defined carotid plaque burden and high-risk plaque composition to subsequent vascular events, although their relative prognostic importance has varied across populations.^30–33^ The PARISK study, for example, identified total plaque volume together with intraplaque hemorrhage as independent predictors of recurrent ipsilateral ischemic stroke or TIA,^30^ whereas other cohorts have demonstrated associations between lipid-rich plaque features and subsequent cardiovascular events.^31–33^ Our study extends these findings providing prospective evidence for the value of assessing plaque morphology with carotid CTA.

### Study limitations

Several limitations should be considered. First, CTA images were acquired using heterogeneous acquisition protocols, and differences in scanner, reconstruction, contrast timing, and image noise may have influenced HU-based plaque classification. Second, participants were pooled from cohorts with different recruitment strategies and potential selection biases, which may limit the generalizability of our findings and may have contributed to an overrepresentation of minor strokes (median NIHSS, 2). Third, the relatively small number of subsequent vascular events limited statistical power and precluded analyses specifically focused on recurrent ipsilateral ischemic stroke; loss to follow-up may have further affected the longitudinal analyses. Fourth, stroke etiology was classified according to TOAST criteria based on diagnostic information available at baseline and was not systematically updated according to subsequent etiologic investigations. Therefore, some patients classified as having an undetermined etiology may have been misclassified, particularly if other potential stroke mechanisms were identified during follow-up.

## Conclusions

Quantitative CTA-based carotid plaque analysis identified greater LRNC volume in plaques ipsilateral to ischemic events of undetermined etiology, an association that persisted after adjustment for wall volume. LRNC was also associated with subsequent vascular risk although the association was attenuated following adjustment for overall carotid plaque burden. Pending further validation, CTA-based assessment of plaque composition and burden may provide complementary information for etiologic assessment and vascular risk stratification.

## Data Availability

The individual-level data underlying this study are not publicly available due to patient privacy, ethical, and institutional data protection restrictions. Requests for access to de-identified data will be directed to the corresponding author and will be considered subject to applicable ethical approvals, data-sharing agreements, and institutional policies.

## Clinical perspectives

### Competency in medical knowledge

Quantitative carotid CTA can characterize plaque burden and vulnerability in patients with ischemic cerebrovascular events, potentially identifying high-risk plaques in strokes of undetermined etiology. In these patients, high-risk plaque features and greater plaque burden are associated with residual vascular risk and may improve risk stratification beyond conventional etiologic classification.

### Translational outlook

Prospective validation and further refinement of CTA-based plaque characterization are needed to establish its value for identifying high-risk plaques, improving etiologic assessment in stroke of undetermined etiology, and refining vascular risk prediction.

## Acknowledgements

We extend our gratitude to all patients and their families for participating in the included studies. We would like to thank the data managers at ISD and all study nurses and physicians for their support with recruitment and data collection.

## Funding Support

The AtherOMICS study and this work were funded by the German Research Foundation’s (DFG) Emmy Noether Programme (GZ GE3461/2-1, ID 512461526 to MKG), the DFG-funded Collaborative Research Center SFB 1744 / CRC 1744 (ID 548585053), the DFG-funded Munich Cluster for Systems Neurology (SyNergy; EXC 2145, ID 390857198 to MKG), the Fritz Thyssen Foundation (Ref. 10.22.2.024 MN to MKG), the Hertie Network of Excellence in Clinical Neuroscience (ID P1230035 to MKG). The DEDEMAS/DEMDAS study was funded by the German Center for Neurodegenerative Diseases (DEMDAS [MC002]). The funders had no role in study design, data collection and analysis, decision to publish, or preparation of the manuscript.

## Author Disclosures

AA serves as a scientific advisor to Elucid Bioimaging. RAP is an employee of Elucid Bioimaging. Both hold stock options in the company. MKG reports consulting fees from Tourmaline bio, Inc., Pheiron GmbH, Dexcel Pharma Technologies Ltd., and Novartis AG all unrelated to this work.

## Abbreviations

CTA: computed tomography angiography
FDR: false discovery rate
LAA: large-artery atherosclerosis
LRNC: lipid-rich necrotic core
NIHSS: National Institutes of Health Stroke Scale
TIA: transient ischemic attack

---

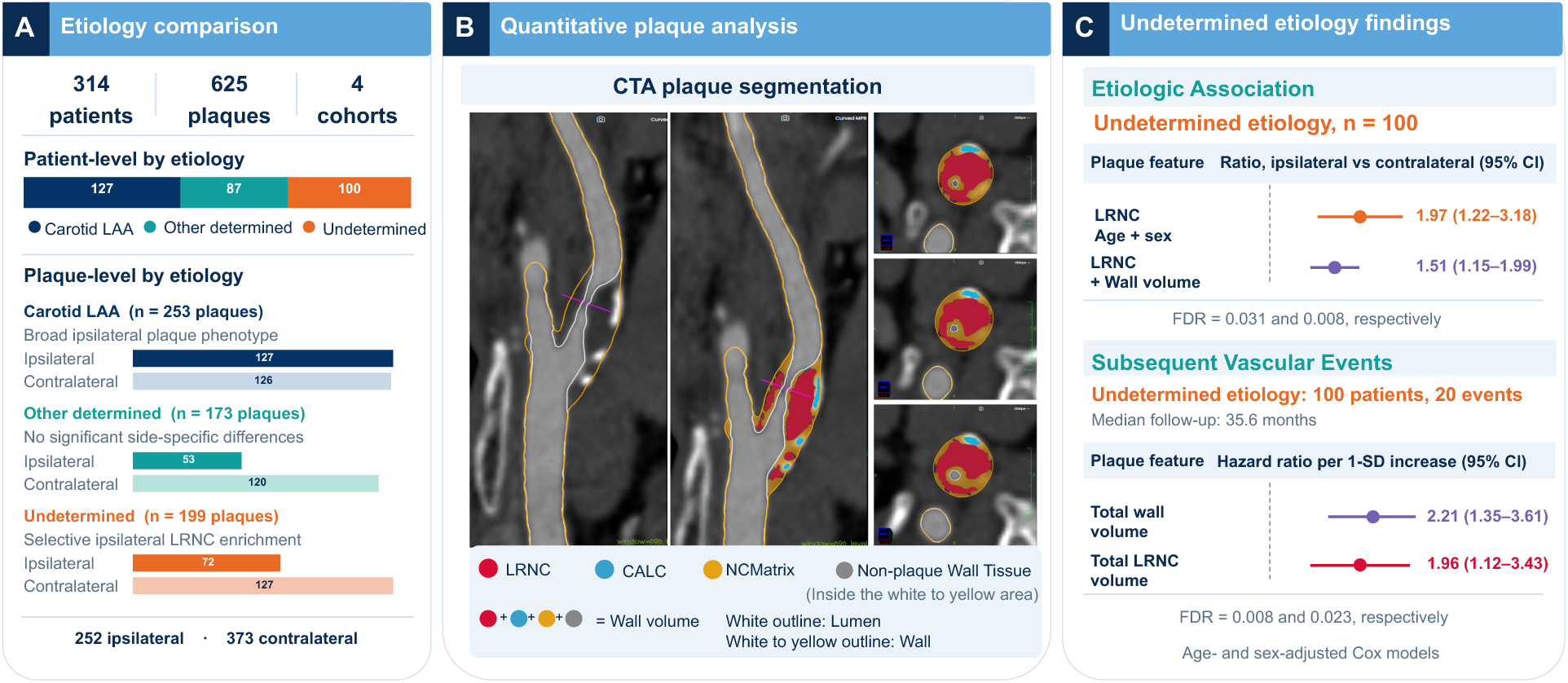
Central Illustration. Carotid Plaque Phenotypes and Vascular Risk in Undetermined Stroke. (A) Distribution of carotid plaques across ischemic stroke etiologies. (B) Quantitative CTA-based plaque segmentation and characterization. (C) In undetermined stroke, ipsilateral LRNC enrichment persisted after adjustment for wall volume, while greater carotid wall and LRNC volumes were associated with subsequent vascular events. Abbreviations: LAA = large-artery atherosclerosis; CTA = computed tomography angiography; LRNC = lipid-rich necrotic core; CALC = calcification; NCMatrix = noncalcified matrix; CI = confidence interval; FDR = false discovery rate.

## Notes

### Clinical Protocols

https://doi.org/10.1126/sciadv.aee2639

### Author Declarations

All participants provided written informed consent for participation in the studies. All studies received ethical approval from the Ethics Committee of the Medical Faculty of LMU Munich: DEDEMAS/DEMDAS (approval numbers 035-11 and 201-13), PROSCIS-M (approval number 366-10), and AtherOMICS (approval numbers 121-09 and 22-0135). The retrospective analysis of the CTA dataset was also approved by the Ethics Committee of the Medical Faculty of LMU Munich (23-0772).

