## Supplementary Material for "Quantitative CTA-Based Carotid Plaque Composition Across Stroke Etiologies and Residual Vascular Risk"

**Supplemental Appendix**

### Supplementary Methods

#### Section S1. Additional study and imaging methods

#### Section S1.1 Source cohorts and study design

Participants in the present study were derived from four parent studies or biobanks conducted at or including LMU University Hospital Munich.

**DEDEMAS**

DEDEMAS[1,2] is a prospective observational study designed to investigate determinants of post-stroke cognitive impairment and dementia after acute stroke. The study collected detailed clinical, neuropsychological, imaging, and biomarker information, with longitudinal follow-up for cognitive and vascular outcomes. For the present analysis, only participants from the Munich site with available cervical CTA and sufficient clinical information were considered.

**DEMDAS**

DEMDAS[3,4] is a longitudinal multicenter observational study investigating vascular and neurodegenerative mechanisms of dementia after stroke. Patients with acute stroke were recruited within the early post-stroke period and underwent repeated clinical, neuropsychological, imaging, and biomarker assessments during long-term follow-up. In the present study, we included only the Munich/ISD subset with analyzable cervical CTA.

**PROSCIS-M**

PROSCIS-M[5] is the Munich arm of the Prospective Cohort with Incident Stroke, a prospective hospital-based cohort of patients with first-ever ischemic stroke. The study was designed to investigate predictors of long-term vascular outcomes, including recurrent stroke, myocardial infarction, vascular death, and all-cause mortality. For the current analysis, patients were selected from the Munich cohort according to the availability and quality of cervical CTA and relevant clinical follow-up information.

**AtherOMICS**

AtherOMICS[6] is a prospective atherosclerosis biobank at LMU Klinikum enrolling patients undergoing carotid or femoral endarterectomy. The study integrates clinical data, preoperative vascular imaging, peripheral blood samples, plaque tissue, and multi-omic profiling. In the present analysis, AtherOMICS participants with available and analyzable cervical CTA and sufficient clinical information were included.

**Section S1.2 CTA acquisition and plaque feature quantification**

CTA examinations were acquired as part of routine acute stroke imaging according to institutional protocols at LMU University Hospital using multidetector CT scanners, including SOMATOM Force, SOMATOM Definition AS+, and SOMATOM Definition Flash systems (Siemens Healthineers, Forchheim, Germany). Image acquisition extended from the aortic arch to the cranial vertex. Iodinated contrast medium was administered intravenously, followed by a saline chaser, both at 5 mL/s. Scanning was initiated using bolus tracking with a threshold of 100 HU in the aortic trunk. Tube voltage was 80 or 120 kVp depending on the scanner, with automated tube-current modulation. Available CTA examinations had reconstructed section thicknesses ranging from 0.6 to 2.5 mm; only examinations with reconstructed section thickness ≤1.5 mm were included in the present analysis.

**Section S1.3 Assessment of future vascular events**

Follow-up procedures differed across the contributing cohorts. In DEDEMAS/DEMDAS, participants were assessed at 3, 6, 12, 24, 36, 48, and 60 months, with comprehensive in-person assessments conducted at 6, 12, 36, and 60 months and interim assessments performed primarily by telephone. In PROSCIS-M, follow-up assessments were conducted over in-person or telephone interviews at 3, 12, 24, and 36 months. Potential outcome events reported by participants or informants were verified through review of medical records, imaging findings, and documentation obtained from treating physicians or hospitals. For the present study, we assessed time to the first occurrence of a composite vascular endpoint. The composite endpoint comprised recurrent stroke, transient ischemic attack, myocardial infarction, angina pectoris, coronary artery disease, or peripheral artery disease during follow-up. Follow-up was calculated from the date of CTA to the first occurrence of any component of the composite endpoint or the last available follow-up.

### Section S2. Statistical analysis

All additional and sensitivity analyses were conducted separately for each plaque characteristic within each stroke-etiology group. In the index-event laterality analyses, each plaque characteristic was entered separately as the dependent variable. In the long-term vascular risk analyses, each patient-level plaque characteristic was entered separately as the exposure variable. All plaque-volume variables were transformed using log10(volume + 1) before modeling. For the long-term vascular risk analyses, the transformed plaque measures were additionally standardized within each stroke-etiology group and model-specific analysis sample.

#### Section S2.1 Index-event laterality analyses

Plaque characteristics were analyzed using log10(volume + 1)-transformed values. The primary analysis used age- and sex-adjusted linear mixed-effects models with plaque-side status (ipsilateral vs. contralateral) as the exposure and a participant-specific random intercept to account for correlation between bilateral measurements. Model coefficients were back-transformed and reported as ipsilateral-to-contralateral geometric mean ratios with 95% confidence intervals.

**Additional adjustment analyses.** The primary models were further adjusted separately for lumen volume, wall volume, or conventional vascular risk factors, including body mass index, hypertension, diabetes mellitus, dyslipidemia, and ever smoking.

**Sensitivity analyses.** An age- and sex-adjusted within-participant paired analysis was restricted to patients with evaluable plaque measurements on both ipsilateral and contralateral sides. For each plaque characteristic, the within-participant difference in the corresponding log10-

transformed value between sides was evaluated. In a second sensitivity analysis, the primary linear mixed-effects analysis was repeated after excluding patients with posterior-circulation or bilateral index-event patterns.

**Section S2.2 Long-term vascular risk analyses**

Associations between plaque characteristics and the long-term composite vascular outcome were evaluated using Cox proportional hazards regression. Follow-up time, outcome definitions, and censoring were defined as in the main Methods. In the primary analysis, patient-level bilateral total values were calculated by summing measurements across evaluable carotid plaques; when only one side was evaluable, the available measurement was used. Plaque measures were log10(volume + 1)-transformed and standardized within each stroke-etiology group and model-specific analysis sample. Each plaque characteristic was entered separately into age- and sex-adjusted Cox models, and hazard ratios were reported per 1-standard-deviation increase in the transformed measure.

**Additional adjustment analyses.** The primary models were further adjusted separately for total lumen volume, total wall volume, or conventional vascular risk factors, including body mass index, hypertension, diabetes mellitus, dyslipidemia, and ever smoking.

**Sensitivity analyses.** First, age- and sex-adjusted side-specific Cox models simultaneously included the ipsilateral and contralateral values of the corresponding plaque characteristic; the reported estimate corresponded to the ipsilateral term conditional on the contralateral value. Second, the primary age- and sex-adjusted analysis was repeated after excluding patients with posterior-circulation or bilateral index-event patterns.

**Alternative wall-volume analyses.** Wall volume was additionally evaluated using bilateral total, maximum-side, ipsilateral, and contralateral values in separate age- and sex-adjusted Cox models. Bilateral total wall volume was defined as the sum of eligible side-specific measurements, and maximum-side wall volume as the larger eligible side-specific value. Ipsilateral and contralateral wall volumes were defined according to index-event laterality.

**Section S2.3 False discovery rate correction**

False discovery rate (FDR) was controlled using the Benjamini–Hochberg procedure. Correction was performed separately within each prespecified stroke-etiology group. The family comprised the five quantitative plaque characteristics: lumen volume, wall volume, calcification volume, lipid-rich necrotic core volume, and noncalcified matrix volume. When lumen or wall volume was included as an adjustment covariate, that characteristic was excluded from the corresponding correction family. For the alternative wall-volume analysis, total, maximum-side, ipsilateral, and contralateral wall volume formed a four-test family within each etiology group.

### Supplementary Tables

**Supplementary Table 1. Baseline characteristics of included participants according to contributing cohort**

| Characteristic | Overall included (n=314) | AtherOMICS (n=96) | DEMDAS (n=61) | DEDEMAS (n=35) | PROSCIS-M (n=122) |
|---|---|---|---|---|---|
| Age, years | 73.0 ± 9.7 | 73.9 ± 9.3 | 72.8 ± 8.5 | 73.0 ± 7.4 | 72.4 ± 11.1 |
| Female sex | 165/314 (52.5%) | 27/96 (28.1%) | 47/61 (77.0%) | 19/35 (54.3%) | 72/122 (59.0%) |
| BMI, kg/m² | 26.5 ± 4.1 | 26.5 ± 4.1 | 27.4 ± 3.6 | 26.0 ± 3.5 | 26.2 ± 4.3 |
| Hypertension | 231/313 (73.8%) | 65/96 (67.7%) | 52/61 (85.2%) | 26/35 (74.3%) | 88/121 (72.7%) |
| Diabetes mellitus | 72/314 (22.9%) | 25/96 (26.0%) | 14/61 (23.0%) | 10/35 (28.6%) | 23/122 (18.9%) |
| Dyslipidemia | 135/304 (44.4%) | 56/86 (65.1%) | 25/61 (41.0%) | 12/35 (34.3%) | 42/122 (34.4%) |
| Ever smoking | 152/314 (48.4%) | 65/96 (67.7%) | 14/61 (23.0%) | 6/35 (17.1%) | 67/122 (54.9%) |
| Coronary artery disease | 64/310 (20.6%) | 30/92 (32.6%) | 7/61 (11.5%) | 5/35 (14.3%) | 22/122 (18.0%) |
| Previous stroke | 45/304 (14.8%) | 31/86 (36.0%) | 9/61 (14.8%) | 5/35 (14.3%) | 0/122 (0.0%) |
| Peripheral artery disease | 26/305 (8.5%) | 16/90 (17.8%) | 1/59 (1.7%) | 1/34 (2.9%) | 8/122 (6.6%) |
| Family history of cardiovascular disease | 163/303 (53.8%) | 46/86 (53.5%) | 33/61 (54.1%) | 16/35 (45.7%) | 68/121 (56.2%) |
| Baseline NIHSS score | 2.0 (1.0–4.0) | 1.0 (0.0–2.2) | 2.0 (1.0–4.0) | 3.0 (1.0–5.8) | 3.0 (1.0–5.0) |

Values are mean ± SD, median (IQR), or n/N (%). No between-cohort comparisons were performed.

**Supplementary Table 2. Baseline characteristics of participants included in and excluded from the present analyses**

| Characteristic | Included (n=314) | Excluded (n=749) | P value |
|---|---|---|---|
| Age, years | 73.0 ± 9.7 | 69.1 ± 11.9 | <0.001 |
| Female sex | 165/314 (52.5%) | 409/747 (54.8%) | 0.511 |
| BMI, kg/m² | 26.5 ± 4.1 | 26.4 ± 5.7 | 0.681 |
| Hypertension | 231/313 (73.8%) | 513/746 (68.8%) | 0.102 |
| Diabetes mellitus | 72/314 (22.9%) | 129/746 (17.3%) | 0.033 |
| Dyslipidemia | 135/304 (44.4%) | 301/735 (41.0%) | 0.305 |
| Ever smoking | 152/314 (48.4%) | 350/747 (46.9%) | 0.644 |
| Coronary artery disease | 64/310 (20.6%) | 145/743 (19.5%) | 0.591 |
| Previous stroke | 45/304 (14.8%) | 57/727 (7.8%) | <0.001 |
| Peripheral artery disease | 26/305 (8.5%) | 83/739 (11.2%) | 0.193 |
| Family history of cardiovascular disease | 163/303 (53.8%) | 391/731 (53.5%) | 0.928 |
| Baseline NIHSS score | 2.0 (1.0–4.0) | 2.0 (1.0–5.0) | 0.272 |

Continuous variables are mean ± SD, NIHSS is median (IQR), and categorical variables are n/N (%), where N is the non-missing denominator. P values use Welch's t test, Mann–Whitney U test, or Pearson's chi-square/Fisher's exact test, as appropriate.

**Supplementary Table 3. Additional adjustment models for index-event laterality of quantitative carotid plaque characteristics.**

| Feature | Model | Ratio (95% CI) | LRT p | FDR |
|---|---|---|---|---|
| **Carotid LAA** | | | | |
| Lumen Volume | model1 | Not applicable | | |
| | model2 | 0.80 (0.75-0.86) | <0.001 | <0.001 |
| | model3 | 0.86 (0.81-0.92) | <0.001 | <0.001 |
| Wall Volume | model1 | 1.73 (1.51-1.98) | <0.001 | <0.001 |
| | model2 | Not applicable | | |
| | model3 | 1.53 (1.34-1.76) | <0.001 | <0.001 |
| CALC Volume | model1 | 1.79 (1.32-2.45) | <0.001 | <0.001 |
| | model2 | 1.08 (0.81-1.45) | 0.589 | 0.589 |
| | model3 | 1.78 (1.30-2.44) | <0.001 | <0.001 |
| LRNC Volume | model1 | 4.17 (2.92-5.96) | <0.001 | <0.001 |
| | model2 | 1.54 (1.19-1.98) | 0.001 | 0.002 |
| | model3 | 3.67 (2.57-5.25) | <0.001 | <0.001 |
| NCMatrix Volume | model1 | 2.26 (1.80-2.82) | <0.001 | <0.001 |
| | model2 | 1.07 (0.97-1.17) | 0.172 | 0.230 |
| | model3 | 2.00 (1.59-2.52) | <0.001 | <0.001 |
| **Other determined** | | | | |
| Lumen Volume | model1 | Not applicable | | |
| | model2 | 0.99 (0.94-1.06) | 0.872 | 0.872 |
| | model3 | 1.00 (0.93-1.06) | 0.885 | 0.885 |
| Wall Volume | model1 | 1.01 (0.85-1.19) | 0.917 | 0.917 |
| | model2 | Not applicable | | |
| | model3 | 0.97 (0.81-1.16) | 0.733 | 0.885 |
| CALC Volume | model1 | 1.13 (0.78-1.64) | 0.513 | 0.917 |
| | model2 | 1.12 (0.81-1.56) | 0.496 | 0.706 |
| | model3 | 1.09 (0.74-1.61) | 0.651 | 0.885 |
| LRNC Volume | model1 | 1.11 (0.66-1.90) | 0.688 | 0.917 |
| | model2 | 1.12 (0.79-1.59) | 0.530 | 0.706 |
| | model3 | 1.05 (0.62-1.79) | 0.845 | 0.885 |
| NCMatrix Volume | model1 | 1.22 (0.81-1.83) | 0.343 | 0.917 |
| | model2 | 1.20 (0.93-1.55) | 0.159 | 0.634 |
| | model3 | 1.14 (0.75-1.72) | 0.536 | 0.885 |
| **Undetermined** | | | | |
| Lumen Volume | model1 | Not applicable | | |
| | model2 | 0.92 (0.87-0.97) | 0.002 | 0.007 |
| | model3 | 0.93 (0.88-0.99) | 0.015 | 0.037 |
| Wall Volume | model1 | 1.18 (1.03-1.37) | 0.021 | 0.028 |
| | model2 | Not applicable | | |
| | model3 | 1.11 (0.95-1.29) | 0.178 | 0.222 |
| CALC Volume | model1 | 1.30 (0.91-1.86) | 0.152 | 0.152 |
| | model2 | 1.05 (0.77-1.44) | 0.757 | 0.757 |
| | model3 | 1.21 (0.84-1.73) | 0.311 | 0.311 |
| LRNC Volume | model1 | 2.08 (1.29-3.36) | 0.003 | 0.012 |
| | model2 | 1.51 (1.15-1.99) | 0.004 | 0.008 |
| | model3 | 1.94 (1.21-3.13) | 0.007 | 0.035 |
| NCMatrix Volume | model1 | 1.50 (1.07-2.10) | 0.019 | 0.028 |
| | model2 | 1.18 (0.95-1.45) | 0.134 | 0.179 |
| | model3 | 1.41 (1.01-1.98) | 0.045 | 0.076 |

Associations between plaque-side status and quantitative carotid plaque characteristics were assessed using carotid-side-level linear mixed-effects models with patient-specific random intercepts. The dependent variable was log10(plaque characteristic volume + 1). Regression coefficients comparing ipsilateral with contralateral plaques were backtransformed and reported as geometric mean ratios with 95% confidence intervals. A ratio greater than 1 indicates a larger plaque characteristic on the event side, whereas a ratio less than 1 indicates a smaller value on the ipsilateral side. Model 1 was adjusted for age, sex, and lumen volume; Model 2 was adjusted for age, sex, and wall volume; and Model 3 was adjusted for age, sex, hypertension, diabetes mellitus, dyslipidemia, BMI, and ever smoking. Analyses were stratified by stroke etiology. P values were obtained using likelihood-ratio tests. False discovery rates were controlled using the Benjamini–Hochberg procedure within each stroke-etiology group and adjustment model across the applicable plaque characteristics. “Not applicable” indicates that the evaluated plaque characteristic was also included as an adjustment covariate.

**Supplementary Table 4. Sensitivity analyses of the associations between quantitative carotid plaque characteristics and index-event laterality**

| Stroke etiology | Feature | Analysis | Patients | Ratio (95% CI) | P value | FDR |
|---|---|---|---|---|---|---|
| Carotid LAA | LRNC | Paired analysis | 126 | 3.67 (2.61-5.17) | <0.001 | <0.001 |
| | LRNC | Excluding posterior-circulation/bilateral index events | 127 | 3.68 (2.62-5.16) | <0.001 | <0.001 |
| | CALC | Paired analysis | 126 | 1.70 (1.26-2.30) | <0.001 | <0.001 |
| | CALC | Excluding posterior-circulation/bilateral index events | 127 | 1.71 (1.27-2.31) | <0.001 | <0.001 |
| | NCMatrix | Paired analysis | 126 | 2.01 (1.61-2.50) | <0.001 | <0.001 |
| | NCMatrix | Excluding posterior-circulation/bilateral index events | 127 | 2.01 (1.62-2.50) | <0.001 | <0.001 |
| Other determined | LRNC | Paired analysis | 50 | 1.06 (0.59-1.91) | 0.843 | 0.925 |
| | LRNC | Excluding posterior-circulation/bilateral index events | 51 | 1.04 (0.59-1.84) | 0.891 | 0.891 |
| | CALC | Paired analysis | 50 | 1.16 (0.78-1.74) | 0.461 | 0.925 |
| | CALC | Excluding posterior-circulation/bilateral index events | 51 | 1.14 (0.78-1.69) | 0.496 | 0.891 |
| | NCMatrix | Paired analysis | 50 | 1.13 (0.72-1.78) | 0.594 | 0.925 |
| | NCMatrix | Excluding posterior-circulation/bilateral index events | 51 | 1.07 (0.69-1.68) | 0.760 | 0.891 |
| Undetermined | LRNC | Paired analysis | 56 | 1.89 (1.12-3.18) | 0.020 | 0.050 |
| | LRNC | Excluding posterior-circulation/bilateral index events | 57 | 1.92 (1.15-3.20) | 0.015 | 0.038 |
| | CALC | Paired analysis | 56 | 1.04 (0.71-1.51) | 0.848 | 0.848 |
| | CALC | Excluding posterior-circulation/bilateral index events | 57 | 1.03 (0.71-1.49) | 0.862 | 0.862 |
| | NCMatrix | Paired analysis | 56 | 1.25 (0.89-1.76) | 0.206 | 0.344 |
| | NCMatrix | Excluding posterior-circulation/bilateral index events | 57 | 1.25 (0.90-1.75) | 0.190 | 0.316 |

Index-event laterality ratios compare ipsilateral with contralateral plaque-characteristic volumes; ratios greater than 1 indicate greater volume on the ipsilateral side. The paired analysis was an age- and sex-adjusted within-participant analysis restricted to patients with evaluable measurements on both carotid sides. The exclusion analysis repeated the age- and sex-adjusted linear mixed-effects analysis after excluding patients with posterior-circulation or bilateral index-event patterns, with a participant-specific random intercept accounting for correlation between carotid sides. Plaque volumes were transformed as log10(volume + 1). P values were obtained from paired linear models for the paired analysis and likelihood-ratio tests for the exclusion analysis. For each analysis, Benjamini–Hochberg false discovery rate correction was performed separately within each stroke-etiology group across all five quantitative plaque characteristics: lumen volume, wall volume, calcification volume, lipid-rich necrotic core volume, and noncalcified matrix volume. The displayed FDR values correspond to these five-test correction families. Abbreviations: CALC, calcification; CI, confidence interval; FDR, false discovery rate; LAA, large-artery atherosclerosis; LRNC, lipid-rich necrotic core; NCMatrix, noncalcified matrix.

**Supplementary Table 5. Etiology-stratified Cox regression analyses of patient-level total carotid plaque characteristics and long-term composite vascular events**

| Feature | Model | n/events | HR (95% CI) | P | FDR |
|---|---|---|---|---|---|
| **Carotid LAA** | | | | | |
| Lumen Volume_total | Age + sex adjusted | 31/10 | 0.83 (0.44-1.59) | 0.581 | 0.857 |
| Wall Volume_total | Age + sex adjusted | 31/10 | 0.84 (0.42-1.66) | 0.608 | 0.857 |
| CALC Volume_total | Age + sex adjusted | 31/10 | 0.83 (0.41-1.72) | 0.622 | 0.857 |
| LRNC Volume_total | Age + sex adjusted | 31/10 | 0.90 (0.47-1.70) | 0.739 | 0.857 |
| NCMatrix Volume_total | Age + sex adjusted | 31/10 | 0.94 (0.49-1.80) | 0.857 | 0.857 |
| **Other determined** | | | | | |
| Lumen Volume_total | Age + sex adjusted | 87/15 | 1.72 (0.92-3.21) | 0.090 | 0.449 |
| Wall Volume_total | Age + sex adjusted | 87/15 | 1.33 (0.71-2.49) | 0.378 | 0.919 |
| CALC Volume_total | Age + sex adjusted | 87/15 | 1.13 (0.66-1.94) | 0.647 | 0.919 |
| LRNC Volume_total | Age + sex adjusted | 87/15 | 0.97 (0.56-1.70) | 0.919 | 0.919 |
| NCMatrix Volume_total | Age + sex adjusted | 87/15 | 0.94 (0.56-1.58) | 0.811 | 0.919 |
| **Undetermined** | | | | | |
| Lumen Volume_total | Age + sex adjusted | 100/20 | 0.93 (0.57-1.52) | 0.771 | 0.771 |
| Wall Volume_total | Age + sex adjusted | 100/20 | 2.21 (1.35-3.61) | 0.002 | 0.008 |
| CALC Volume_total | Age + sex adjusted | 100/20 | 2.04 (1.23-3.36) | 0.005 | 0.009 |
| LRNC Volume_total | Age + sex adjusted | 100/20 | 1.96 (1.12-3.43) | 0.019 | 0.023 |
| NCMatrix Volume_total | Age + sex adjusted | 100/20 | 2.41 (1.35-4.33) | 0.003 | 0.008 |

Associations between patient-level total bilateral carotid plaque characteristics and long-term composite vascular events were assessed using Cox proportional-hazards models stratified by stroke etiology. The composite endpoint comprised recurrent stroke, transient ischemic attack, myocardial infarction, angina pectoris, coronary artery disease, or peripheral artery disease during follow-up. For each patient, eligible left- and right-sided measurements were summed. Plaque characteristics were transformed as log10(total bilateral volume + 1) and standardized within each stroke-etiology group and analysis sample. Hazard ratios are reported per 1-standard-deviation increase in the transformed plaque characteristic. All models were adjusted for age and sex. P values were obtained from Wald tests. Benjamini-Hochberg false discovery rate correction was performed within each stroke-etiology group across the five quantitative plaque characteristics. Analyses included patients with positive follow-up time and complete data for the analysis. Abbreviations: CI, confidence interval; FDR, false discovery rate; HR, hazard ratio; LAA, large-artery atherosclerosis; NCMatrix, noncalcified matrix.

**Supplementary Table 6. Additional adjustment models for the associations between quantitative carotid plaque characteristics and long-term composite vascular events.**

| Features | Model | HR (95% CI) | p value | FDR |
|---|---|---|---|---|
| **Carotid LAA total/events: 31/10** | | | | |
| Lumen Volume | model1 | Not applicable | | |
| | model2 | 0.88 (0.42-1.84) | 0.735 | 0.878 |
| | model3 | 1.73 (0.56-5.31) | 0.337 | 0.886 |
| Wall Volume | model1 | 0.89 (0.40-1.96) | 0.768 | 0.925 |
| | model2 | Not applicable | | |
| | model3 | 1.14 (0.51-2.55) | 0.751 | 0.886 |
| CALC Volume | model1 | 0.90 (0.38-2.11) | 0.807 | 0.925 |
| | model2 | 0.89 (0.39-2.05) | 0.782 | 0.878 |
| | model3 | 0.94 (0.38-2.32) | 0.886 | 0.886 |
| LRNC Volume | model1 | 0.92 (0.46-1.84) | 0.814 | 0.925 |
| | model2 | 1.09 (0.35-3.40) | 0.878 | 0.878 |
| | model3 | 0.88 (0.36-2.12) | 0.774 | 0.886 |
| NCMatrix Volume | model1 | 0.97 (0.48-1.94) | 0.925 | 0.925 |
| | model2 | 1.63 (0.31-8.67) | 0.564 | 0.878 |
| | model3 | 1.16 (0.46-2.88) | 0.754 | 0.886 |
| **Other determined total/events: 87/15** | | | | |
| Lumen Volume | model1 | Not applicable | | |
| | model2 | 1.69 (0.85-3.36) | 0.132 | 0.263 |
| | model3 | 1.71 (0.89-3.31) | 0.110 | 0.549 |
| Wall Volume | model1 | 1.04 (0.49-2.21) | 0.921 | 0.921 |
| | model2 | Not applicable | | |
| | model3 | 1.21 (0.62-2.35) | 0.581 | 0.902 |
| CALC Volume | model1 | 1.03 (0.62-1.71) | 0.907 | 0.921 |
| | model2 | 1.04 (0.61-1.77) | 0.880 | 0.880 |
| | model3 | 1.03 (0.60-1.77) | 0.902 | 0.902 |
| LRNC Volume | model1 | 0.92 (0.50-1.67) | 0.773 | 0.921 |
| | model2 | 0.69 (0.33-1.47) | 0.340 | 0.454 |
| | model3 | 0.91 (0.50-1.65) | 0.746 | 0.902 |
| NCMatrix Volume | model1 | 0.72 (0.40-1.33) | 0.296 | 0.921 |
| | model2 | 0.36 (0.15-0.82) | 0.016 | 0.064 |
| | model3 | 0.83 (0.47-1.46) | 0.515 | 0.902 |
| **Undetermined total/events: 100/20** | | | | |
| Lumen Volume | model1 | Not applicable | | |
| | model2 | 0.60 (0.35-1.03) | 0.066 | 0.264 |
| | model3 | 0.79 (0.46-1.36) | 0.396 | 0.396 |
| Wall Volume | model1 | 2.64 (1.56-4.47) | <0.001 | 0.001 |
| | model2 | Not applicable | | |
| | model3 | 2.10 (1.24-3.56) | 0.006 | 0.017 |
| CALC Volume | model1 | 2.24 (1.31-3.83) | 0.003 | 0.004 |
| | model2 | 1.36 (0.72-2.54) | 0.342 | 0.637 |
| | model3 | 2.23 (1.21-4.12) | 0.010 | 0.017 |
| LRNC Volume | model1 | 2.07 (1.15-3.72) | 0.015 | 0.015 |
| | model2 | 0.80 (0.31-2.06) | 0.637 | 0.637 |
| | model3 | 1.87 (1.04-3.35) | 0.035 | 0.044 |
| NCMatrix Volume | model1 | 2.74 (1.46-5.13) | 0.002 | 0.003 |
| | model2 | 1.44 (0.36-5.81) | 0.612 | 0.637 |
| | model3 | 2.18 (1.22-3.91) | 0.009 | 0.017 |

Associations between patient-level total bilateral carotid plaque characteristics and long-term composite vascular events were assessed using Cox proportional-hazards models stratified by stroke etiology. The composite endpoint comprised recurrent stroke, transient ischemic attack, myocardial infarction, angina pectoris, coronary artery disease, or peripheral artery disease during follow-up Plaque characteristics were calculated as the sum of all eligible carotid-side measurements for each patient, transformed as log10(total bilateral volume + 1), and standardized within each stroke-etiology group and model-analysis sample. Hazard ratios are reported per 1-standard-deviation increase in the transformed plaque characteristic. Total lumen and wall volume covariates were entered on the log10(total bilateral volume + 1) scale. Model 1 was adjusted for age, sex, and total lumen volume; Model 2 was adjusted for age, sex, and total wall volume; and Model 3 was adjusted for age, sex, hypertension, diabetes mellitus, dyslipidemia, BMI, and ever smoking. P values were obtained from Wald tests. False discovery rates were controlled using the Benjamini–Hochberg procedure within each stroke-etiology group and adjustment model across applicable plaque characteristics. "Not applicable" indicates that the evaluated plaque characteristic was also included as an adjustment covariate.

**Supplementary Table 7. Sensitivity analyses of the associations between quantitative carotid plaque characteristics and long-term vascular risk**

| Stroke etiology | Plaque characteristic | Analysis | HR (95% CI) | P value | FDR |
|---|---|---|---|---|---|
| Carotid LAA | LRNC | Side-specific Cox analysis | 0.88 (0.44-1.78) | 0.728 | 0.911 |
| | LRNC | Excluding posterior-circulation/bilateral index events | 0.90 (0.47-1.70) | 0.739 | 0.857 |
| | CALC | Side-specific Cox analysis | 0.36 (0.12-1.04) | 0.058 | 0.292 |
| | CALC | Excluding posterior-circulation/bilateral index events | 0.83 (0.41-1.72) | 0.622 | 0.857 |
| | NCMatrix | Side-specific Cox analysis | 0.79 (0.39-1.59) | 0.501 | 0.835 |
| | NCMatrix | Excluding posterior-circulation/bilateral index events | 0.94 (0.49-1.80) | 0.857 | 0.857 |
| Other determined | LRNC | Side-specific Cox analysis | 1.51 (0.60-3.79) | 0.383 | 0.602 |
| | LRNC | Excluding posterior-circulation/bilateral index events | 0.94 (0.40-2.19) | 0.881 | 0.881 |
| | CALC | Side-specific Cox analysis | 1.67 (0.56-5.01) | 0.356 | 0.602 |
| | CALC | Excluding posterior-circulation/bilateral index events | 1.89 (0.63-5.66) | 0.253 | 0.634 |
| | NCMatrix | Side-specific Cox analysis | 1.49 (0.49-4.52) | 0.482 | 0.602 |
| | NCMatrix | Excluding posterior-circulation/bilateral index events | 1.08 (0.43-2.71) | 0.875 | 0.881 |
| Undetermined | LRNC | Side-specific Cox analysis | 2.03 (0.91-4.54) | 0.085 | 0.142 |
| | LRNC | Excluding posterior-circulation/bilateral index events | 1.96 (0.90-4.27) | 0.089 | 0.112 |
| | CALC | Side-specific Cox analysis | 1.64 (0.74-3.65) | 0.222 | 0.278 |
| | CALC | Excluding posterior-circulation/bilateral index events | 2.13 (1.03-4.42) | 0.042 | 0.070 |
| | NCMatrix | Side-specific Cox analysis | 2.68 (1.00-7.16) | 0.050 | 0.126 |
| | NCMatrix | Excluding posterior-circulation/bilateral index events | 2.60 (1.09-6.20) | 0.031 | 0.070 |

Associations with long-term vascular risk were assessed using age- and sex-adjusted Cox proportional hazards regression. In the side-specific analysis, ipsilateral and contralateral values of the corresponding plaque characteristic were entered simultaneously into the Cox model, and the reported HR corresponds to the ipsilateral term conditional on the contralateral value. In the exclusion analysis, patients with posterior-circulation or bilateral index-event patterns were excluded, and the patient-level bilateral plaque measure was defined as the sum of eligible left- and right-sided measurements. Plaque measures were transformed as log10(volume + 1), and the exposure corresponding to each reported HR was standardized within each stroke-etiology group and model-specific analysis sample. HRs are reported per 1-SD increase in the transformed plaque measure. The composite endpoint comprised recurrent stroke, transient ischemic attack, myocardial infarction, angina pectoris, coronary artery disease, or peripheral artery disease during follow-up. P values were obtained from Wald tests. Benjamini–Hochberg false discovery rate correction was performed separately within each stroke-etiology group across the corresponding ipsilateral coefficients for all five quantitative plaque characteristics in the side-specific analysis and across all five quantitative plaque characteristics in the exclusion analysis. Abbreviations: CALC, calcification; CI, confidence interval; FDR, false discovery rate; HR, hazard ratio; LAA, large-artery atherosclerosis; LRNC, lipid-rich necrotic core; NCMatrix, noncalcified matrix; SD, standard deviation.

**Supplementary Table 8. Etiology-stratified Cox regression analyses of alternative wall-volume summaries and long-term composite vascular events**

| Feature | Model | n/events | HR (95% CI) | P | FDR |
|---|---|---|---|---|---|
| Carotid LAA | | | | | |
| Wall Volume_total | Age + sex adjusted | 31/10 | 0.84 (0.42-1.66) | 0.608 | 0.841 |
| Wall Volume_max | Age + sex adjusted | 31/10 | 0.85 (0.44-1.64) | 0.631 | 0.841 |
| Wall Volume_ipsilateral | Age + sex adjusted | 31/10 | 0.74 (0.36-1.53) | 0.423 | 0.841 |
| Wall Volume_contralateral | Age + sex adjusted | 31/10 | 0.98 (0.51-1.91) | 0.964 | 0.964 |
| Other determined | | | | | |
| Wall Volume_total | Age + sex adjusted | 87/15 | 1.33 (0.71-2.49) | 0.378 | 0.448 |
| Wall Volume_max | Age + sex adjusted | 87/15 | 1.29 (0.70-2.38) | 0.415 | 0.448 |
| Wall Volume_ipsilateral | Age + sex adjusted | 52/7 | 1.80 (0.53-6.09) | 0.346 | 0.448 |
| Wall Volume_contralateral | Age + sex adjusted | 85/15 | 1.27 (0.69-2.35) | 0.448 | 0.448 |
| Undetermined | | | | | |
| Wall Volume_total | Age + sex adjusted | 100/20 | 2.21 (1.35-3.61) | 0.002 | 0.003 |
| Wall Volume_max | Age + sex adjusted | 100/20 | 2.23 (1.37-3.63) | 0.001 | 0.003 |
| Wall Volume_ipsilateral | Age + sex adjusted | 64/16 | 2.20 (1.32-3.66) | 0.003 | 0.003 |
| Wall Volume_contralateral | Age + sex adjusted | 92/15 | 1.82 (1.04-3.18) | 0.035 | 0.035 |

Associations between alternative patient-level wall-volume summaries and long-term composite vascular events were assessed using age- and sex-adjusted Cox proportional hazards regression stratified by stroke etiology. The composite endpoint comprised recurrent stroke, transient ischemic attack, myocardial infarction, angina pectoris, coronary artery disease, or peripheral artery disease during follow-up. Total wall volume was defined as the sum of eligible left- and right-sided measurements; maximum wall volume as the larger eligible side-specific value; and ipsilateral and contralateral wall volumes according to index-event laterality. Wall-volume summaries were log10(volume + 1)-transformed and standardized within each stroke-etiology group and model-specific analysis sample. HRs are reported per 1-SD increase in the transformed wall-volume summary. P values were obtained from Wald tests. FDR was controlled using the Benjamini–Hochberg procedure separately within each stroke-etiology group. Analyses included patients with positive follow-up time and complete data for the corresponding model. Abbreviations: CI, confidence interval; FDR, false discovery rate; HR, hazard ratio; LAA, large-artery atherosclerosis; SD, standard deviation; TIA, transient ischemic attack.

**Supplementary Table 9. Distribution of first composite vascular events during follow-up according to index stroke etiology.**

| First vascular event during follow-up | Carotid LAA (N=31) | Other determined (N=87) | Undetermined (N=100) | Overall (N=218) |
|---|---|---|---|---|
| Any composite vascular event | 10 (32.3%) | 15 (17.2%) | 20 (20.0%) | 45 (20.6%) |
| Recurrent stroke | 1 (3.2%) | 4 (4.6%) | 9 (9.0%) | 14 (6.4%) |
| Transient ischemic attack | 2 (6.5%) | 3 (3.4%) | 3 (3.0%) | 8 (3.7%) |
| Myocardial infarction | 2 (6.5%) | 2 (2.3%) | 1 (1.0%) | 5 (2.3%) |
| Angina pectoris | 1 (3.2%) | 1 (1.1%) | 1 (1.0%) | 3 (1.4%) |
| Coronary artery disease | 3 (9.7%) | 3 (3.4%) | 3 (3.0%) | 9 (4.1%) |
| Peripheral artery disease | 1 (3.2%) | 2 (2.3%) | 3 (3.0%) | 6 (2.8%) |
| No composite vascular event during follow-up | 21 (67.7%) | 72 (82.8%) | 80 (80.0%) | 173 (79.4%) |
| Patients included in the longitudinal analysis | 31 (100.0%) | 87 (100.0%) | 100 (100.0%) | 218 (100.0%) |

Values are presented as n (%), with percentages calculated using the total number of patients in each stroke-etiology group as the denominator. Stroke etiologies were classified as carotid LAA, other determined etiology, or undetermined etiology. The composite vascular endpoint comprised recurrent stroke, transient ischemic attack, myocardial infarction, angina pectoris, coronary artery disease, or peripheral artery disease. Only the first qualifying vascular event during follow-up was counted for each patient. Patients without a composite vascular event were censored at their last available follow-up. Abbreviations: LAA, large-artery atherosclerosis.
